# Seroconversion panels demonstrate anti-SARS-CoV-2 antibody development after administration of the mRNA-1273 vaccine

**DOI:** 10.1101/2021.06.20.21258152

**Authors:** Francisco Belda, Oscar Mora, Monica Lopez-Martinez, Nerea Torres, Ana Vivanco, Rebecca Christie, Michael Crowley

**Affiliations:** Research and Development, Bio Supplies Division, Grifols, 08174 Sant Cugat del Vallès (Barcelona), Spain; R&D Department, Progenika Biopharma, A Grifols Company, Ibaizabal Bidea, Edificio 504, Parque Tecnológico de Bizkaia, 48160 Derio, Bizkaia, Spain; Access Biologicals, Vista, CA 92081, USA

**Keywords:** COVID-19, SARS-CoV-2, mRNA-1273 vaccine, immunoglobulin, immunity, seroconversion panel

## Abstract

Seroconversion panels are an important tool for investigating antibody responses in acute and chronic phases of disease and development of serological assays for viral diseases including COVID-19. Globally it is anticipated that vaccines against SARS-CoV-2 will facilitate control of the current pandemic. The two COVID-19 seroconversion panels analyzed in this study were obtained from consenting donors with samples collected before vaccination with the mRNA-1273 vaccine (Moderna) and after the first and second doses of the vaccine. Panel samples were tested for antibodies to SARS-CoV-2 (IgG). Individual subjects with a positive response for anti-SARS-CoV2 IgG in their pre-vaccination samples showed a significantly enhanced response to the first vaccination. In older subjects, weaker immunological responses to the first injection were observed, which were overcome by the second injection. All subjects in the study were positive for anti-SARS-CoV-2 IgG after the second dose of vaccine.

## 1. Introduction

Since December 2019, COVID-19, the disease resulting from SARS-CoV-2 infection, has affected more than 148 million people worldwide and resulted in at least 3.1 million deaths. [1] Containment of COVID-19 to date has depended heavily on public health measures such as testing and tracing, mask wearing, social/physical distancing, and hand washing. Treatments available to date have not been completely effective and have relied on anti-inflammatory steroids (primarily dexamethasone) [2, 3], antivirals (remdesivir) [4–6] and passive immunity (e.g., convalescent plasma [7–9] and monoclonal antibodies such as bamlanivimab and etesevimab [10–12]). Recent development of vaccines against SARS-CoV-2 [13, 14] has provided the healthcare community with crucial tools that can ultimately bring the current pandemic under control. In this paper, the development of seroconversion panels with samples collected before and after administration (two doses) of a SARS-CoV-2 vaccine are described to demonstrate the immune response to schedule doses of the vaccine.

Seroconversion panels are a series of blood samples collected before and after the development of antibodies in response to a viral infection or vaccine. These panels can be useful tools in the creation of antibody assays, determination of the window period of detection, validation and quality control for development and manufacture of commercial antibody tests. Detection of viral exposure and vaccine effectiveness through immunologic assays is a critical step in gaining control of SARS-CoV-2 and returning to normalcy. The panels are also a source of well-defined (prior and after vaccination) neutralizing antibodies useful in investigating their effectiveness blocking the new COVID-19 variants.

The COVID-19 seroconversion panels analyzed in this study were collected from two groups of subjects with samples collected before vaccination (mRNA-1273 SARS-CoV-2 vaccine, Moderna, Cambridge, MA, USA) and after the first and second doses of the vaccine. The samples were analyzed using a chemiluminescent immunoassay (CLIA) and an enzyme-linked immunosorbent assay (ELISA).

## 2. Material & Methods

The samples which make up these seroconversion panels were collected from consenting donors at a hospital in Tennessee (USA). The samples were collected with informed consent under an approved IRB protocol ([1149706-4] Diagnostic QC and Pre-Clinical Sample Collection Project: Ballad Health System Institutional Review Board, Johnson City, TN, USA) and in compliance with all applicable regulatory guidelines.

Two seroconversion panels were used in this study to characterize the appearance of anti-SARS-CoV-2 IgG after administration of the mRNA-1273 SARS-CoV-2 vaccine: 15 subjects ere included in COVID-19 Vaccine Panel G and 30 subjects were included in COVID-19 Vaccine Panel H (Access Biologicals, Vista, CA, USA). For both panels, samples were collected prior to the first vaccination (objective target ≤ 2 days: sample 1), prior to the second vaccination (objective target ≤ 2 days: sample 2), and after the second vaccination (objective target 13-15 days; sample 3). The samples collected for these seroconversion panels were either serum samples collected in serum separating tubes (Panels G and H) and/or plasma samples collected in the presence of potassium EDTA (Panel H). Samples were stored at −20° C until use. The samples were thawed at room temperature and gently mixed by inversion prior to testing.

Panel G was comprised of undiluted, unpreserved serum specimens collected from 15 subjects between 22nd December 2020 and 25th February 2021. The subjects were healthy adults 21 to 76 years of age who received two injections of mRNA-1273 SARS-CoV-2 vaccine (100 µg) objective target 28 days apart. There were 5 male and 10 female subjects in this group. All the subjects were Caucasian.

As noted above, the samples in Panel G were all serum divided into 1 mL aliquots. The samples in Panel G were tested with a CLIA (Liaison SARS-CoV-2 IgG Assay, Diasorin, Inc, Saluggia, Italy: EUA approved) and an ELISA (Progenika anti-SARS-CoV-2 IgG kit, Progenika Biopharma, Derio, Bizkaia, Spain: CE-IVD certified immunoassay).

Seroconversion COVID-19 Vaccine Panel H was comprised of undiluted, unpreserved serum and potassium EDTA-treated plasma specimens collected from 30 subjects between 23rd December 2020 to 15th March 2021. The subjects were healthy adults 19 to 73 years of age who received two injections of mRNA-1273 SARS-CoV-2 vaccine (100 µg) objective target 28 days apart. They were 9 male and 21 female subjects in this panel. The subjects were 29 Caucasians and one African American. Samples from this panel were tested using the CLIA described above (Diasorin).

The CLIA and ELISA SARS-CoV-2 IgG assays were performed according to the manufacturers’ directions. Both assays utilize recombinant antigens specific to SARS-CoV-2. The Diasorin assay uses antigens to S1 and S2 spike protein IgGs, while the Progenika assay uses antigens to S1 spike protein IgG. The specific composition of the antigens was not specified but they were developed independently and are presumed to be nonidentical.

Quantitative variables were compared using Mann-Whitney-Wilcoxon (JMP software 16.0).

The vaccine administered to the subjects in Panel G and Panel H was the mRNA-1273 SARS-CoV-2 vaccine (Moderna, Cambridge, MA, USA), a lipid nanoparticle-encapsulated mRNA vaccine. The mRNA in this vaccine encodes for the perfusion-stabilized full-length SARS-CoV-2 spike protein. This vaccine has been proven to be highly efficacious in a phase 3 randomized, observer-blinded, placebo-controlled clinical trial – preventing 94% of Covid-19 illness in treated subjects compared to the placebo group. [13]

## 3. Results

Testing of pre-vaccination samples in Panel G (n=15 subjects) with the CLIA gave the following results (Figure 1: Front Row): 13 negative, 2 positive results (Subjects 5 and 15). The ELISA gave similar results (Figure 2: Front Row): 12 negative, 2 positive (Subjects 5 and 15) and 1 equivocal result (Subject 10). The presence of anti-SARS-CoV-2 IgG in these two subjects indicates that they were previously infected with the virus prior to collection of these samples.

**Figure 1.**
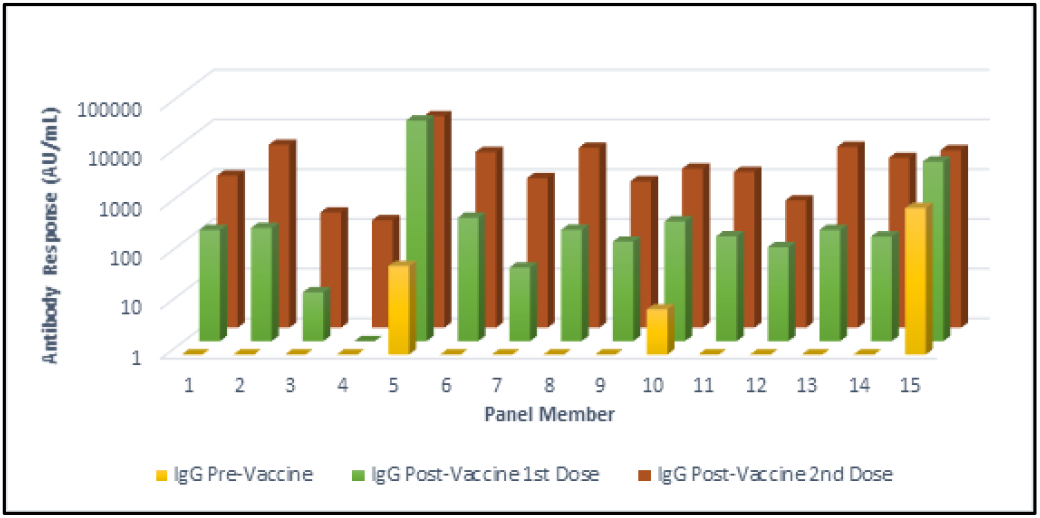
Antibody responses in a seroconversion panel (Panel G) of 15 subjects before and after vaccination with two doses of the mRNA-1273 SARS-CoV-2 vaccine (Moderna, Cambridge, MA, USA). These results were obtained using a chemiluminescent immunoassay. (AU/mL) Arbitrary Unit. Responses ≥ 15.0 units were considered positive. The front row shows results from samples collected prior to the first vaccination. The middle row shows the results from samples collected after the first vaccination and prior to the second vaccination. The back row shows the results from samples collected after the second vaccination.

**Figure 2.**
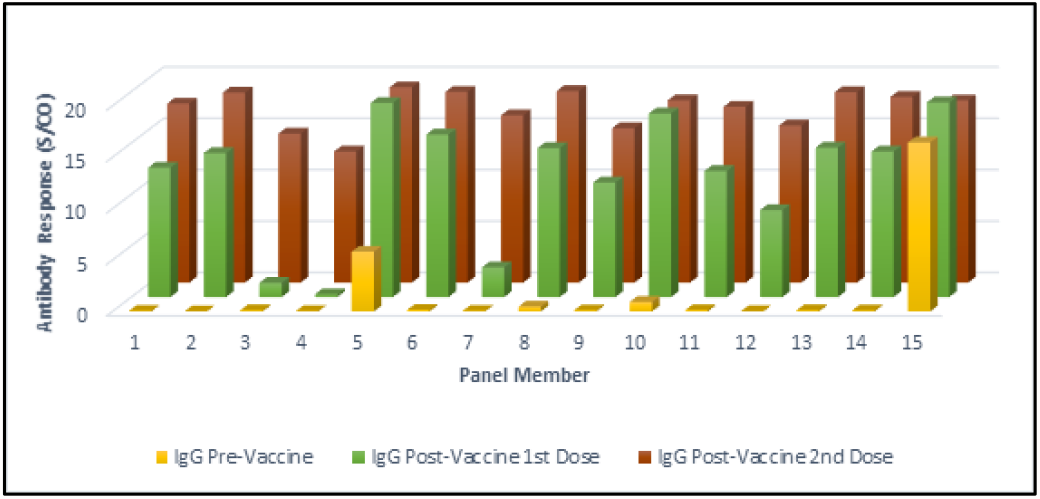
Antibody responses in a seroconversion panel (Panel G) of 15 subjects before and after vaccination with two doses of the mRNA-1273 SARS-CoV-2 vaccine (Moderna, Cambridge, MA, USA). These results were obtained using an enzyme-linked immunosorbent assay (ELISA). (S/CO) Signal-to-cutoff values. Responses ≥ 1.1 S/CO were considered positive. The front row shows the results from samples collected prior to the first vaccination. The middle row shows the results from samples collected after the first vaccination and prior to the second vaccination. The back row shows the results from samples collected after the second vaccination.

The samples in Panel G collected after the first injection of the mRNA-1273 vaccine showed 13 positive and 2 negative results using the CLIA (Figure 1: Middle Row). The ELISA showed 14 positive and 1 negative result after the first vaccine injection (Figure 2: Middle Row). The highest antibody responses detected after the first injection in both assays were in the two subjects who had detectable antibodies in their pre-injection samples (Subjects 5 and 15). The negative samples were collected from two of the oldest subjects in the panel (Subjects 3 and 4: >70 years old).

Samples collected after the second injection of the mRNA-1273 vaccine were positive in all 15 subjects in both assays (Figure 1: Back Row and Figure 2: Back Row). Overall, there was good agreement between the results of the CLIA and the ELISA assays.

With Panel H (n=30 subjects), the pre-vaccine samples showed 24 negatives and 6 positives results (Figure 3: Front Row). Subject 19 was near the cut-off value and was counted as negative. The six positives pre-vaccination samples were seen with Subjects 2, 8, 12, 16, 24 and 30. The samples in Panel H were tested using only the SARS-CoV-2 IgG CLIA (Diasorin).

**Figure 3.**
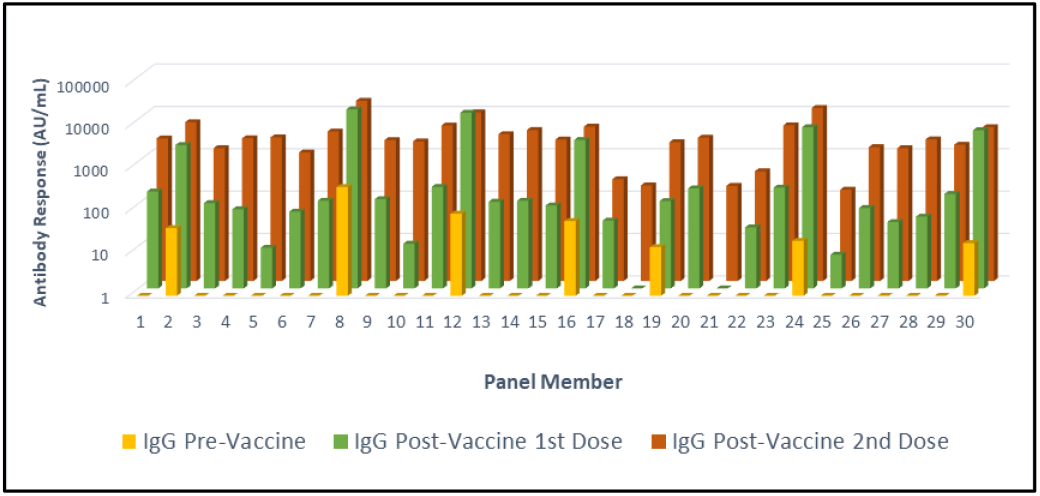
Antibody responses in a seroconversion panel (Panel H) of 30 subjects before and after vaccination with two doses of the mRNA-1273 SARS-CoV-2 vaccine (Moderna, Cambridge, MA, USA). These results were obtained using a chemiluminescent immunoassay. (AU/mL) Arbitrary Unit. Responses ≥ 15.0 (AU/mL) were considered positive. The front row shows the results from samples collected prior to the first vaccination. The middle row shows the results from samples collected after the first vaccination and prior to the second vaccination. The back row shows the results from samples collected after the second vaccination.

The samples collected after the first vaccine dose showed 25 positive and 5 negative results. (Figure 3: Middle Row). Six of these positive samples (samples from Subjects 2, 8, 12, 16, 24 and 30) showed a marked increase in IgG values and corresponded to subjects with positive pre-vaccination samples. These positive pre-vaccination samples indicate previous COVID infections. The negative samples were collected from two of the oldest subjects in the panel (Subjects 18 and 21: > 70 years old) and the other three from people 55-70 years old (Subjects 5, 10 and 25).

The samples in Panel H collected after the second vaccine dose were positive for all 30 subjects (Figure 3: Back Row).

Figure 4 shows the immune response from all subjects in Panels G and H including the pre-vaccination, post first dose and post second dose samples.

**Figure 4.**
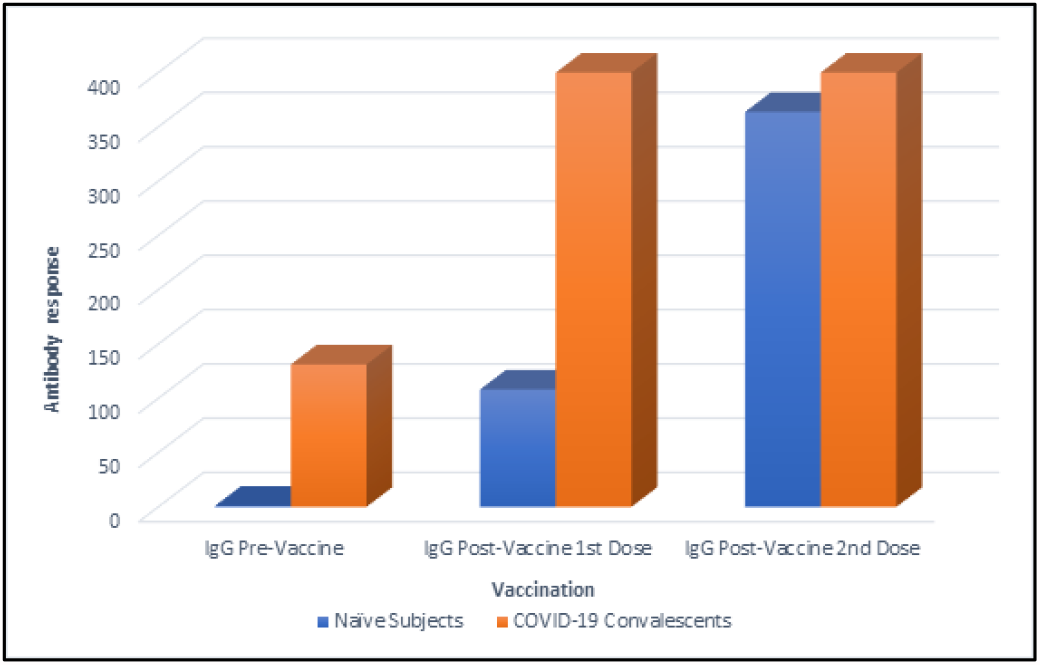
Comparison of antibody responses between naïve subject versus COVID-19 convalescent subjects prior to and after vaccination. Antibody responses were measured in samples collected prior to the first vaccination, after the first vaccination and prior to the second vaccination and after the second vaccination with the mRNA-1273 SARS-CoV-2 vaccine (Moderna, Cambridge, MA, USA). These results were obtained using a chemiluminescent immunoassay. (AU/mL) Arbitrary Unit. Responses ≥ 15.0 (AU/mL) were considered positive; values greater than the assay range (up to 400 AU/ml) were recorded as 400 AU/ml.

The data are grouped into naïve subjects (no immunologic response in pre-vaccination samples) and COVID-19 convalescent subjects (positive response for anti-S1/S2 Ig G prior to vaccination: mean ± SD; 131.2 ± 157.9 arbitrary units (AU/ml)). Samples collected after the first vaccine showed significantly higher Ig G levels (> 400 AU/ml vs 108.1 ± 82.8 AU/ml; p<0.0001) in the convalescent subject samples compared to naïve subjects. Immunological responses after the second vaccine dose were similar, no significant differences were observed between both groups (363.7 ± 81.8 AU/ml in naïve subjects and > 400 AU/ml in the convalescent subjects; p=0.154).

## 4. Discussion

The measurement of IgG levels has been and will continue to be a crucial tool in monitoring and controlling the spread of COVID-19 and in the determination of medication and vaccine effectiveness. Detection of antibodies by immunoassay is useful for the verification of humoral immunity whether due to past infection, passive immunity through administration of convalescent plasma or monoclonal antibodies, or vaccination.

A seroconversion panel can be used for the creation, development and production of antibody immunoassays. In fact, some regulatory authorities have issued guidelines requiring or recommending the use of seroconversion panels in the validation of serological assays for detection of viral infection. [15] Seroconversion panels are available for several viral diseases: hepatitis A, hepatitis B, hepatitis C, and human immunodeficiency virus (HIV).

A seroconversion panel of samples from a single subject who had community-acquired COVID-19 has been previously described. [16] The previously described seroconversion panel was the result of detection of SARS-CoV-2 infection in a regular plasma donor. This frequent donor had several pre-infection samples available from previous donations. Pre-infection samples are not available for most COVID-19 patients as blood is usually not collected until treatment is initiated.

Because the seroconversion panels described in this paper were centered around vaccination, pre-conversion samples were available for most of these subjects. There were a few subjects in each panel (n=2 for Panel G and n=6 for Panel H) that were positive for anti-SARS-CoV-2 IgG prior to vaccination. These results indicate these subjects had exposure and convalescence to COVID-19 prior to collection of these pre-vaccination samples. Subjects that showed a positive result in their pre-vaccination samples had the highest levels of IgG after the first vaccine dose. These results are consistent with the observation of greater antibody responses to a first vaccine dose in patients with a history of COVID-19. [17–19] This increased response to the first vaccine has been linked to an increased level of SARS-CoV-2 antigen specific memory B cells. [17]

The antibody response to the first vaccine dose in subjects that showed a positive result in their pre-vaccination samples (convalescent subjects) was of the same magnitude as that obtained with the second vaccine dose in subjects without previous COVID-19 infection (naïve subjects). This supports the idea of prioritizing vaccine administration based on the serostatus during the current global vaccine shortage, thus maximizing coverage and effectiveness of current supplies. [18 - 20]

Panel G was analyzed with both a CLIA and an ELISA. These two tests gave similar results indicating that these seroconversion panels could be useful for performing comparisons of different immunoassays to determine which is the most useful for an application.

Limitations of this study include the number of subjects studied and a racial distribution that is not a representative of the total population. Another limitation is that the detection assays used are not neutralizing antibodies assays that will detect the actual amount of viral blocking antibodies. Furthermore, it would be interesting to determine the presence of specific antibodies against different antigenic parts of the virus than S1 and S2 spike proteins (e.g. nucleocapside protein), in the group of post-vaccination samples from COVID-19 convalescent subjects.

In conclusion, the data presented here show that these seroconversion panels demonstrate the appearance of anti-SARS-CoV-2 IgG after vaccination. The data also show that individual subjects with previous COVID-19 show an enhanced response to the first vaccination. It was also observed that weaker responses to the first injection – observed in four elderly subjects (over 70 years old) and three younger subjects (55-70 years old) – were overcome by the second injection. All subjects in these seroconversion panels generated anti-SARS-CoV-2 IgG in response to the mRNA-1273 SARS-CoV-2 vaccine. These data show that these conversion panels could be useful tools for the development and comparison of serological tests for COVID-19 and quality control during their manufacture.

## Data Availability

All the relevant data that support the findings of this study are available within the article. Complementary data can be available from the corresponding author upon reasonable request

## Acknowledgements

Michael K. James, PhD is acknowledged for medical writing and Jordi Bozzo, PhD, CMPP for editorial assistance. Contributions from Norbert Piel, Sansan Lin, Rodrigo Gajardo and Jerry A. Holmberg (Grifols) who provided their expert opinions are also acknowledged.

## Conflict of Interest

Francisco Belda, Oscar Mora, Monica Lopez Martinez, Nerea Torres, and Ana Vivanco are employees of Grifols. Rebecca Christie and Michael Crowley are employees of Access Biologicals..

## Funding

These studies were supported by Grifols (Barcelona, Spain) and Access Biologicals (Vista, CA, USA).

## Notes

### Author Declarations

The samples were collected with informed consent under an approved IRB protocol ([1149706-4] Diagnostic QC and Pre-Clinical Sample Collection Project: Ballad Health System Institutional Review Board, Johnson City, TN, USA) and in compliance with all applicable regulatory guidelines.

## References

[1] Johns Hopkins University Center for Systems Science and Engineering. COVID-19 Dashboard. Baltimore, MD: Johns Hopkins University Coronavirus Resource Center. Accessed 28 April 2021.

[2] Recovery Collaborative Group, Horby P, Lim WS, Emberson JR, Mafham M, Bell JL, et al. Dexamethasone in Hospitalized Patients with Covid-19. N Engl J Med. 2021;384:693–704.

[3] WHO Rapid Evidence Appraisal for COVID-19 Therapies Working Group, Sterne JAC, Murthy S, Diaz JV, Slutsky AS, Villar J, et al. Association Between Administration of Systemic Corticosteroids and Mortality Among Critically Ill Patients With COVID-19: A Meta-analysis. JAMA. 2020;324:1330–41.

[4] Beigel JH, Tomashek KM, Dodd LE, Mehta AK, Zingman BS, Kalil AC, et al. Remdesivir for the Treatment of Covid-19 - Final Report. N Engl J Med. 2020;383:1813–26.

[5] Goldman JD, Lye DCB, Hui DS, Marks KM, Bruno R, Montejano R, et al. Remdesivir for 5 or 10 Days in Patients with Severe Covid-19. N Engl J Med. 2020;383:1827–37.

[6] Spinner CD, Gottlieb RL, Criner GJ, Arribas Lopez JR, Cattelan AM, Soriano Viladomiu A, et al. Effect of Remdesivir vs Standard Care on Clinical Status at 11 Days in Patients With Moderate COVID-19: A Randomized Clinical Trial. JAMA. 2020;324:1048–57.

[7] Li L, Zhang W, Hu Y, Tong X, Zheng S, Yang J, et al. Effect of Convalescent Plasma Therapy on Time to Clinical Improvement in Patients With Severe and Life-threatening COVID-19: A Randomized Clinical Trial. JAMA. 2020;324:460–70.

[8] Simonovich VA, Burgos Pratx LD, Scibona P, Beruto MV, Vallone MG, Vazquez C, et al. A Randomized Trial of Convalescent Plasma in Covid-19 Severe Pneumonia. N Engl J Med. 2020.

[9] Shen C, Wang Z, Zhao F, Yang Y, Li J, Yuan J, et al. Treatment of 5 Critically Ill Patients With COVID-19 With Convalescent Plasma. JAMA. 2020;323:1582–9.

[10] Weinreich DM, Sivapalasingam S, Norton T, Ali S, Gao H, Bhore R, et al. REGN-COV2, a Neutralizing Antibody Cocktail, in Outpatients with Covid-19. N Engl J Med. 2021;384:238–51.

[11] Chen P, Nirula A, Heller B, Gottlieb RL, Boscia J, Morris J, et al. SARS-CoV-2 Neutralizing Antibody LY-CoV555 in Outpatients with Covid-19. N Engl J Med. 2021;384:229–37.

[12] Gottlieb RL, Nirula A, Chen P, Boscia J, Heller B, Morris J, et al. Effect of Bamlanivimab as Monotherapy or in Combination With Etesevimab on Viral Load in Patients With Mild to Moderate COVID-19: A Randomized Clinical Trial. JAMA. 2021;325:632–44.

[13] Baden LR, El Sahly HM, Essink B, Kotloff K, Frey S, Novak R, et al. Efficacy and Safety of the mRNA-1273 SARS-CoV-2 Vaccine. N Engl J Med. 2021;384:403–16.

[14] Walsh EE, Frenck RW, Falsey AR, Kitchin N, Absalon J, Gurtman A, et al. Safety and Immunogenicity of Two RNA-Based Covid-19 Vaccine Candidates. New England Journal of Medicine. 2020.

[15] Food and Drug Administration. Guidance for industry and FDA Staff: Class II special controls guidance document: Hepatitis A serological assays. US Food and Drug Administration, Center for Devices and Radiological Health, Bethesda, MD, USA: US Department of Health and Human Services; 2006. vailable at: https://www.fda.gov/media/71388/download. Accessed 6 July 2020.

[16] Belda F, Lopez-Martinez M, Torres N, Cherenzia R, Crowley M. Available COVID-19 serial seroconversion panel for validation of SARS-CoV-2 antibody assays. Diagn Microbiol Infect Dis. 2021;100:115340.

[17] Goel RR, Apostolidis SA, Painter MM, Mathew D, Pattekar A, Kuthuru O, et al. Distinct antibody and memory B cell responses in SARS-CoV-2 naïve and recovered individuals following mRNA vaccination. Science Immunology. 2021;6:eabi6950.

[18] Azzi L, Focosi D, Dentali F, Baj A, Maggi F. Anti-SARS-CoV-2 RBD IgG responses in convalescent versus naive BNT162b2 vaccine recipients. Vaccine. 2021;39:2489–90.

[19] Mazzoni A, Di Lauria N, Maggi L, Salvati L, Vanni A, Capone M, et al. First-dose mRNA vaccination is sufficient to reactivate immunological memory to SARS-CoV-2 in recovered COVID-19 subjects. J Clin Invest. 2021.

[20] Levi R, Azzolini E, Pozzi C, Ubaldi L, Lagioia M, Mantovani A, and Rescigno M. One dose of SARS-CoV-2 vaccines exponentially increases antibodies in recovered individuals with symptomatic COVID-19. J Clin Invest. 2021. https://doi.org/10.1172/JCI149154.

